# Volumetric Analysis of the Deltoid Muscle From Neck-to-Knee Dixon MRI: A pilot population assessment of a posterior intramuscular injection target

**DOI:** 10.64898/2026.11.10.26367545

**Authors:** Marili Niglas, Nicolas Basty, Fatemeh Dashti, Madhur Varadpande, Marjola Thanaj, Andrew R Davies, Brandon Whitcher, Jędrzej Krawczyk, Jimmy David Bell, E. Louise Thomas, Dimitri Amiras

## Abstract

**Background:** Deltoid muscle volume varies across anatomical regions and population groups, with potential implications for intramuscular injection site selection. Evidence comparing middle and posterior deltoid volumes in vaccine-eligible populations is limited.

**Purpose:** To compare middle and posterior deltoid muscle volumes by vaccine eligibility group, sex, and side.

**Methods:** Deltoid muscles were manually segmented into anterior, middle and posterior regions in 43 UK Biobank Dixon MRI scans. A MONAI deep learning model trained on the annotations (Dice score = 0.78) segmented deltoid muscles of 5,239 participants, categorised by UK flu vaccination eligibility: A (≥65 years without risk conditions, n=1,760), B (≥65 with risk conditions, n=1,744), and C (<65 with risk conditions, n=1,735). Bilateral-average posterior and middle volumes were compared using a paired t-test. A linear mixed-effects model assessed posterior-middle differences by group, sex and side.

**Results:** Mean age was 66.7 ± 7.2 years and body mass index was 27.5 ± 5.7 kg/m^2^ (mean ± SD). Posterior deltoid volume exceeded the middle deltoid volume by 55.36 ml (95% CI, 54.14-56.58, p<0.0001). Posterior-middle differences were largest in Group A, followed by B and then C, greater in men, and greater on the right side (all p<0.0001). Mean segmentation-quality score was 4.27 ± 0.73 on a five-point Likert scale and weighted inter-observer kappa was 0.583 (95% CI, 0.452 - 0.713).

**Conclusions:** Among individuals eligible for the UK flu immunisation, posterior deltoid volume was greater than middle deltoid volume. These findings support further anatomical assessment of the posterior deltoid as a potentially safer injection target.

**Key Results:**

- The deltoid muscles of 5,239 participants from the UK Biobank matching UK annual influenza vaccination eligibility were segmented using a deep learning-based model, enabling precise volumetric quantification of the anterior, middle and posterior deltoids.
- The posterior deltoid demonstrated a larger average muscle volume when compared to the middle volume across vaccine-eligible groups and sexes (p<0.0001).
- These findings provide a comprehensive reference framework for deltoid assessment across different ages and genders, enhancing both research and clinical decision-making for IM injection site selection.

**Summary statement:** Automated deltoid volumetry in a pilot UK Biobank cohort revealed greater posterior than middle deltoid volume, supporting further evaluation of the posterior deltoid for a potential alternative intramuscular vaccination site.

## Introduction

Shoulder Injury Related to Vaccine Administration (SIRVA) encompasses a spectrum of adverse events following immunisation that arise from presumed improper intramuscular (IM) injection into the deltoid region [1]. First described in 2007, Atanasoff et al. reported a series of 13 cases of persistent shoulder pain and dysfunction after deltoid vaccination, establishing SIRVA as a medicolegal entity [2]. Commonly associated pathologies include bursitis, rotator cuff tears, adhesive capsulitis, glenohumeral synovitis, atraumatic osteonecrosis of the humeral head, chondral injury, and axillary or radial nerve injuries [3, 4]. Although the precise pathogenesis of SIRVA remains incompletely understood, it has been proposed that an immune-mediated inflammatory response could be triggered by inadvertent deposition of antigenic material into synovial tissues.

In response, safety guidelines specifying needle length and injection technique have been established [5]. Current recommendations position the injection site two to three fingerbreadths (≈ 5 centimetres) below the acromion process, targeting the central portion of the middle deltoid to avoid neurovascular injury [6, 7]. However, research has shown that this injection site has the potential to damage the subacromial subdeltoid bursa and/or anterior branch of the axillary nerve [6, 8, 9]. Despite landmarking techniques proposed to identify the thickest point of the deltoid muscle, there is limited evidence supporting their efficacy [4].

Earlier studies focused on determining appropriate needle length for the adult deltoid (IM) injections [10-14]. Contrary to current recommendations [13], these investigations suggested different needle-length protocols to improve vaccine delivery and reduce SIRVA risk [10-12]. Yet, their applicability is constrained by small cohorts, heterogeneous populations, and anatomical variability, highlighting the need for more reliable landmarking [11, 12, 14]. Notably, only 54% of trained immunisers correctly identified the recommended injection zone on standardised anatomical diagrams, highlighting the difficulty of current landmarking techniques [4]. More recently, Mardourian et al. [12]proposed an alternative injection site, located more posterior and inferior to the current guidelines, 4 cm distal to the posterolateral acromion corner, effectively targeting the posterior deltoid. However, systematic anatomical evaluation of alternative IM injection targets, particularly in populations who experience annual vaccinations, remains scarce.

Given these limitations, we hypothesised that the deltoid muscle volume would differ between the middle and posterior regions across vaccine-eligible groups. To investigate this, we developed an automated deltoid segmentation model and quantified deltoid muscle volumetric profiles using MRI scans from the UK Biobank.

## Materials and Methods

### Data

The UK Biobank is a large prospective population-based cohort comprising over 500,000 participants, aged 40 to 85, recruited across the United Kingdom between 2006 and 2010. The present study used 5,333 MRI scans available for deltoid processing at the time of the analysis. UK Biobank received ethical approval from the North West Multicentre Research Ethics Committee (REC reference: 11/NW/0382), and all participants provided written informed consent. Access to the UK Biobank resource was granted through an approved health-related research application (Application 23889). All analyses were conducted in accordance with relevant institutional guidelines and regulations.

### Phenotype definitions

Demographic, anthropometric, lifestyle, and physiological measurements were obtained at the UK Biobank imaging visit. These included age, sex, height, weight, waist and hip circumference, dominant-hand grip strength (HGS), metabolic equivalent task (MET), total muscle volume and iliopsoas muscle volume. BMI and waist-to-hip ratio were derived from these measurements. Ethnicity was defined using self-reported ethnic background recorded at baseline assessment, and socioeconomic status was assessed using the Townsend deprivation index calculated immediately prior to the initial assessment visit.

Participants were stratified into three groups based on the UK national flu immunisation programme (NFIP) eligibility criteria [15]: Group A - individuals aged >= 65 years without clinical risk conditions, Group B - participants aged >= 65 years with clinical risk conditions, and Group C – participants aged < 65 years with clinical risk conditions. All conditions were identified using International Classification of Diseases (ICD-9 and ICD-10) codes, Operating Procedure Codes Supplement (OPCS), and relevant self-reported fields available from the UK Biobank. A detailed list of included conditions and the relevant ICD-9 and ICD-10 codes is provided in ***Supplementary Table 1***.

### Image processing and analysis

We utilised the 3D two-point Dixon sequences, which give coverage from neck to knees, from 5,333 participants representative of the previously described immunisation groups in the UK Biobank. Comprehensive details on the MRI abdominal protocol have been reported previously [16]. The six Dixon neckl⍰tol⍰knee MRI series, acquired on a 1.5T scanner (Siemens Healthineers, Erlangen, Germany), were resampled, aligned, corrected for bias-field inhomogeneities, and blended into a single neck-to-knee volume (matrix: 224×174×370; resolution: 2.232×2.232×3.0^3^ mm ) using an automated preprocessing pipeline [17, 18].

### Manual Annotation

Images from 43 UK Biobank participants were manually annotated by MV and supervised by DA (with 13 years of specialist musculoskeletal radiological experience). Both the left and right deltoid muscles were annotated. The anterior medial border of the deltoid was defined by the deltopectoral groove, posteriorly, the border was defined by the posterior scapular and infraspinatus. For each manually annotated deltoid, landmarks were also placed at the anterior and posterior lateral corners of the acromion and along the humeral shaft. Hence, a total of six landmarks were placed in each shoulder: anterior corner of the acromion, posterior corner of the acromion, centre of the humeral head, centre of the humerus inferiorly, most inferior border of the anterior and middle deltoids, and most inferior border of the middle and posterior deltoids. The landmarks were used to construct two planes along the humeral shaft, dividing the deltoid into anterior, middle, and posterior parts. The overview of the landmarks, planes and deltoid subregions can be viewed in ***Figure 1***. The resulting multilabel masks were subsequently checked and validated by DA.

**Figure 1.**
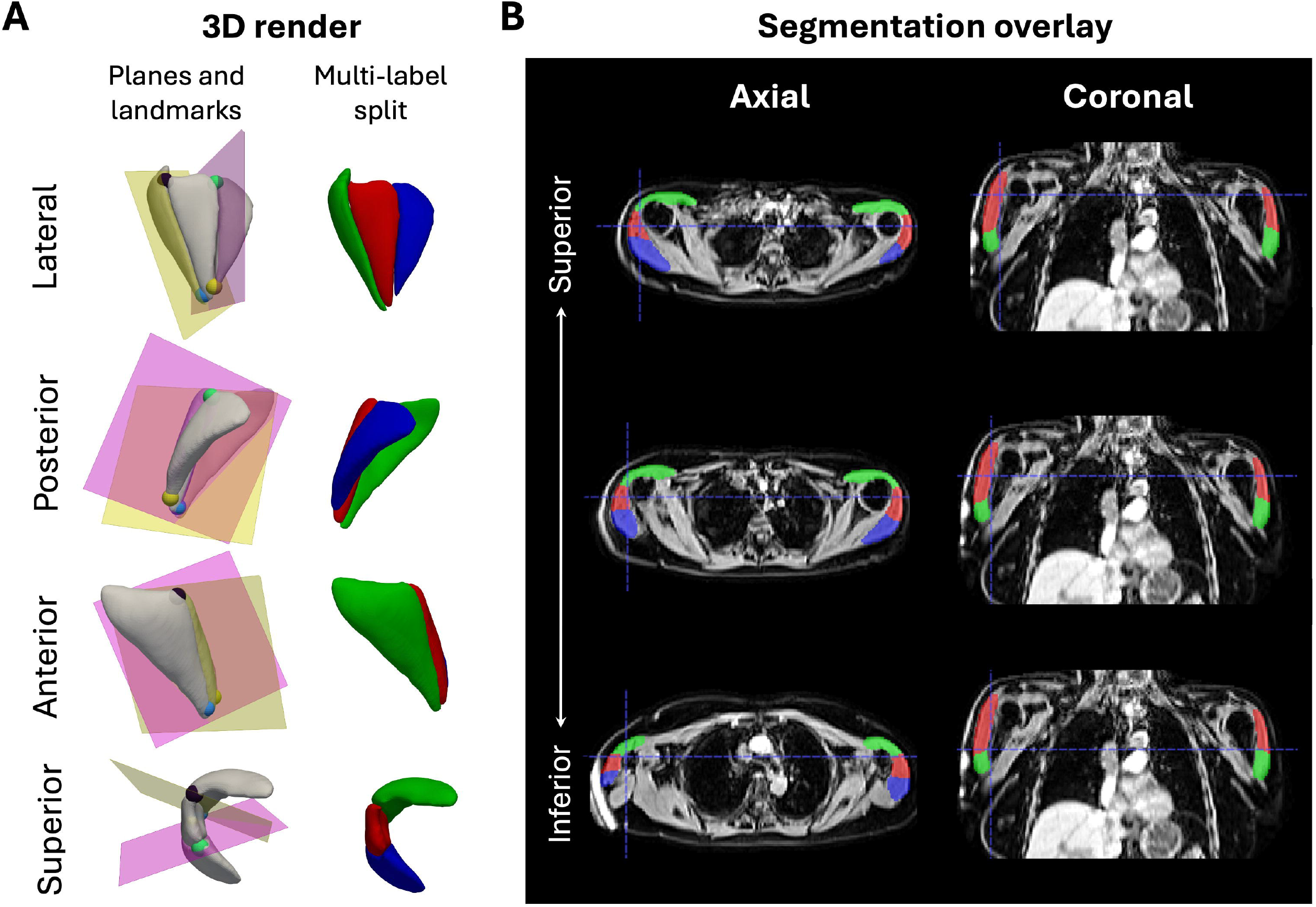
Representative dataset showing the automated segmentation of the deltoid subregions. (A) 3D renders of the deltoid landmark placements, based on the acromial margins, subsequent plane intersections with the deltoid muscle segmentation, and final multi-label deltoid muscle splitting into the anterior, middle, and posterior parts from the lateral, posterior, anterior, and superior views. (B) Dixon water channel image with the automated anterior (green), middle (red), and posterior (blue) deltoid muscle masks as an overlay in axial and coronal planes. Image reproduced by kind permission of UK Biobank ©.

### Segmentation Model

A deep learning segmentation model was trained on the 43 manually annotated multilabel deltoid masks using a 90:10 split of training and test data. The segmentation model produced three tissue classes: anterior, middle, and posterior deltoid muscle. The left and right shoulders were separated in post-processing, resulting in six labels per participant. The Swin-UNETR model architecture was selected based on its strong performance in segmenting complex multi-label medical imaging data [19]. Implementation was performed using MONAI [20] with weights from self-supervised pre-training datasets [21]. Model performance on out-of-sample data achieved a Dice coefficient of 0.78, indicating reliable segmentation accuracy across deltoid parts.

### Quality control

Quality control procedures identified participants with extremely low deltoid muscle volumes, defined separately by sex as values >3 SD below the average normalised deltoid volume. All flagged cases were visually reviewed. These outliers were primarily due to partial inclusion of the deltoid in the field-of-view, rather than segmentation errors (***Supplementary Figure 1***). In total, 94 participants, 33 men and 61 women, were excluded from final analysis, resulting in a participant cohort size of 5,239. No further exclusions were required for missing anthropometric, biological or lifestyle factor variables.

To assess the validity of the automated segmentation method, two independent observers (MV and DA) reviewed the deltoid muscle masks and assigned quality scores using a 5-point Likert scale, based on [22], (1 = poor to 5 = excellent quality) for each muscle compartment (anterior, middle, posterior) on both sides. Inter-observer agreement was reported as mean ± SD, and agreement was quantified using weighted Cohen’s kappa with quadratic weights as well as using the exact percentage of agreement.

### Statistical analysis

All summary statistics and hypothesis tests were conducted in the R statistical computing environment, version 4.6.1 [23]. Deltoid muscle volumes in millilitres were extracted for the anterior, middle, and posterior parts of both sides. Continuous variables were expressed as mean ± standard deviation (SD), and categorical characteristics as counts (percentage). Any differences between volumes were reported as means with 95% confidence intervals.

A paired t-test was used to compare the bilateral-averaged posterior and middle deltoid volumes. Posterior-middle differences were calculated separately for each side and analysed using a linear mixed-effects model. Vaccination-eligibility group, sex, and side were included as fixed effects, and participant as the random intercept to account for the same participants’ left-right measurement non-independence. Post-hoc pairwise group comparisons between vaccine-eligibility groups used Tukey adjustment. A two-sided p<0.05 was considered statistically significant.

## Results

### Cohort characteristics

A total of 5,239 participants were included, of whom 49.7% were women: 1,760 in Group A, 1,744 in Group B, and 1,735 in Group C (as previously defined). The overall mean age of participants was 66.67 ± 7.22 years, and body mass index was 27.54 ± 5.70 kg/m^2.^Most participants self-identified as white (98%). Group C was younger than Groups A and B and had the highest mean body mass index, total muscle volume, and iliopsoas muscle volume. Additional demographic, anthropometric, and activity measurements are presented descriptively in ***Table 1***.

**Table 1.** Table of population characteristics. The table reports the anthropometric and lifestyle variables for the overall cohort, as well as for groups A, B and C, where the continuous variables are presented as mean ± standard deviation and categorical variables as counts (proportion %). No between-group hypothesis tests were performed.

### Segmentation quality

The segmentation quality was considered good, as the overall 5-point Likert scale segmentation-quality score was 4.27 ± 0.73. Scores for individual parts ranged from 3.88 ± 0.72 for the right posterior part to 4.65 ± 0.48 for the right anterior part. Across 120 paired ratings from 20 participants, weighted kappa was 0.583 (95% CI, 0.452-0.713) and exact agreement was 62.5%. Results for each deltoid part are shown in ***Table 2***.

**Table 2.** Segmentation quality and inter-observer agreement. Twenty participants, selected at random with stratification by sex and vaccination-eligibility group, were rated for six compartments. Likert scores range from 1 (poor) to 5 (excellent) and are pooled across both readers. Weighted kappa is reported with its analytic 95% confidence interval. Overall agreement pools 120 paired ratings.

### Deltoid muscle quantification

The mean total deltoid volume was 260.99 ± 59.76 ml. Total deltoid volume was greater in men (307.16 ± 41.04 versus 214.35 ± 33.88 ml) and on the right (275.85 ± 72.30 versus 246.14 ± 68.87 ml). Across vaccination-eligibility groups, total deltoid volume was 263.36 ± 58.36 ml in Group A, 255.37 ± 56.59 ml in Group B, and 264.25 ± 63.76 ml in Group C.

The posterior deltoid muscle volume was the largest anatomical sub-region overall (111.88 ± 39.83 ml), followed by anterior (92.59 ± 19.78 ml) and middle (56.52 ± 17.69 ml) deltoid. The same ordering was seen in women and men, across all three vaccination-eligibility groups, and on both sides (***Table 3***). The posterior bilateral averaged deltoid volume was significantly larger than the middle deltoid volume. The mean posterior-middle deltoid volume difference was 55.36 ml (95% CI 54.14-56.58, p<0.0001) (***Figure 2***).

**Table 3.** Middle, anterior and posterior deltoid volumes reported per group for the whole cohort of 5,333 participants. Values are mean ± SD in ml. Each participant contributes the average of left and right volumes.

**Figure 2.**
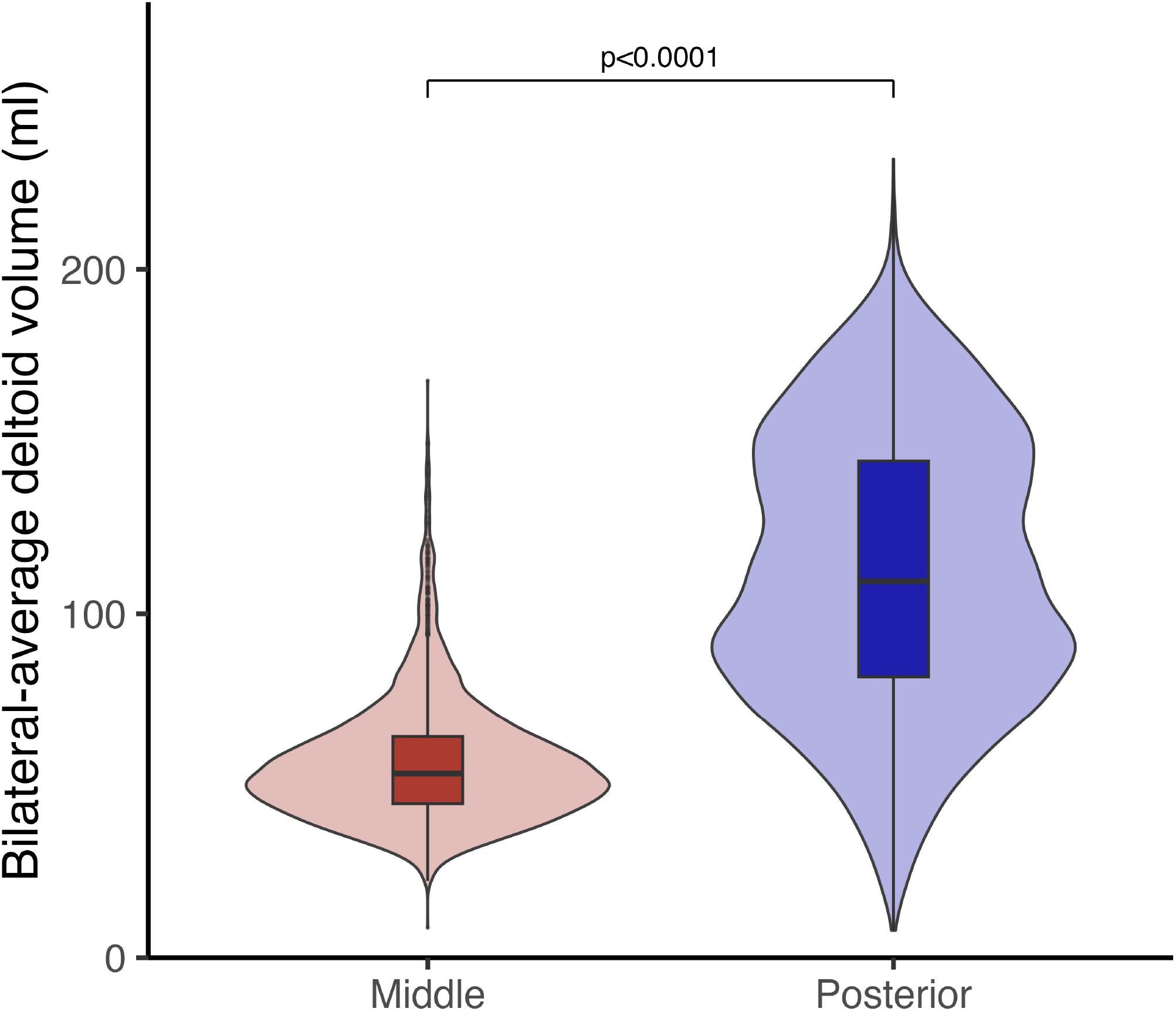
Comparison of middle and posterior deltoid volumes. Violin plots show the distributions of bilateral-average middle and posterior deltoid volumes, with boxplots showing the median and interquartile range. Each participant contributed one bilateral-average value for each deltoid region. The p-value was derived from a paired t-test between bilateral averages of middle and posterior deltoids.

The posterior-middle deltoid volume difference varied by vaccination-eligibility group, sex, and side (all p<0.001). The posterior-middle difference was 63.96 ml (95% CI, 61.85 - 66.07) in Group A, 55.38 ml (53.26 - 57.50) in Group B, and 46.23 ml (44.10 - 48.35) in Group C (***Table 4, Figure 3***). Relative to the corresponding mean total deltoid volume, the posterior-middle volume difference represented approximately 24.3%, 21.7% and 17.5% differences in Groups A, B, and C, respectively. Group C therefore had the highest total deltoid volume despite having the smallest posterior-middle difference. In post-hoc pairwise comparisons, the posterior-middle difference was 8.58 ml greater in Group A than Group B, 17.73 ml greater in Group A than Group C, and 9.15 ml greater in Group B than Group C (all Tukey-adjusted p<0.001, ***Table S1***).

**Table 4.** Posterior-minus-middle deltoid volume differences. Estimates are adjusted for the other model terms. Group and sex estimates are averaged across sides, side estimates are averaged across groups and sex. Each p-value is the overall test of the corresponding model term. Pairwise group comparisons are reported in Table S2.

**Figure 3.**
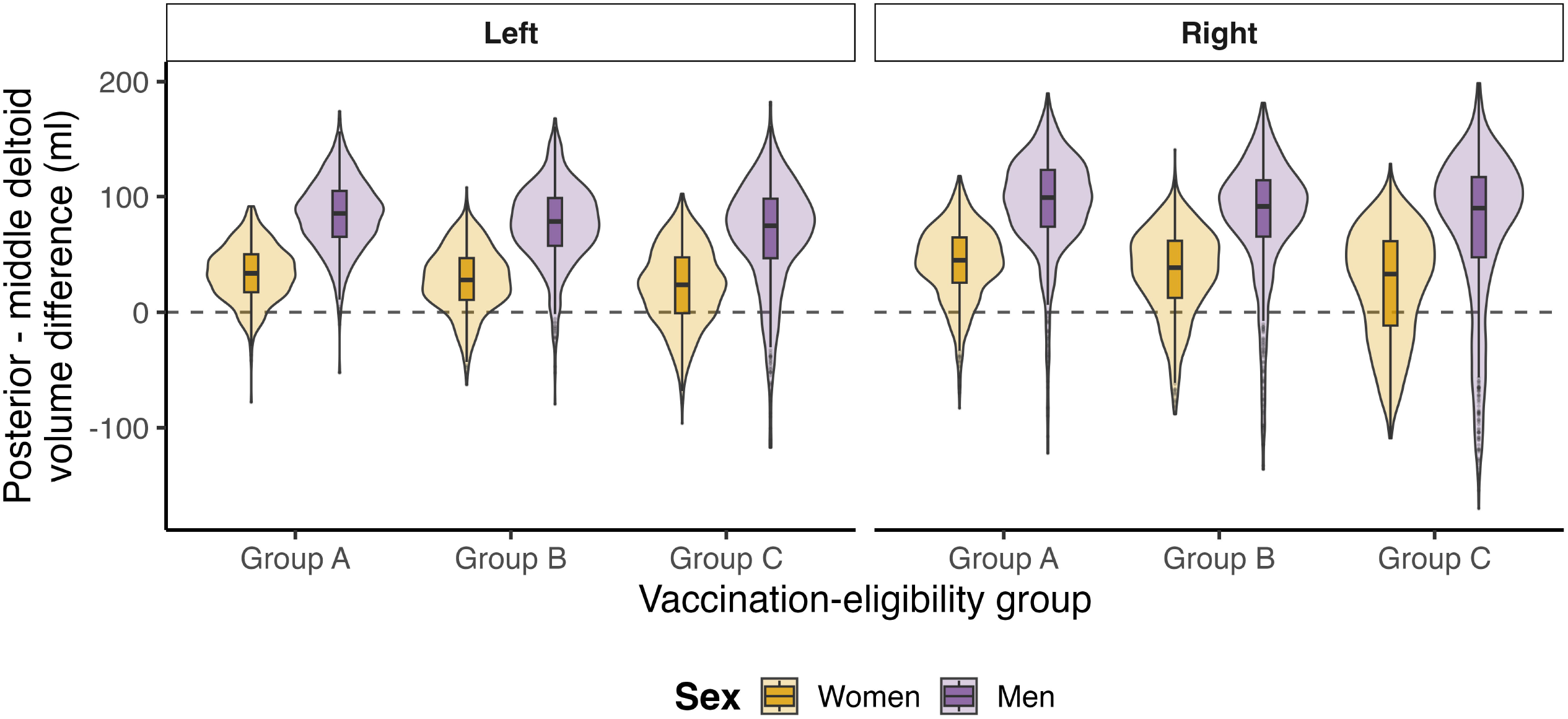
Posterior-middle deltoid volume differences by vaccination-eligibility group, sex, and side. Violin plots show the observed distributions of the posterior-middle volume difference for women and men in each vaccination-eligibility group, separated by left and right side. Boxplots show the median and interquartile range. Positive values indicate a greater posterior than middle deltoid volume, and the dashed horizontal line indicates no difference.

**Figure 4.**
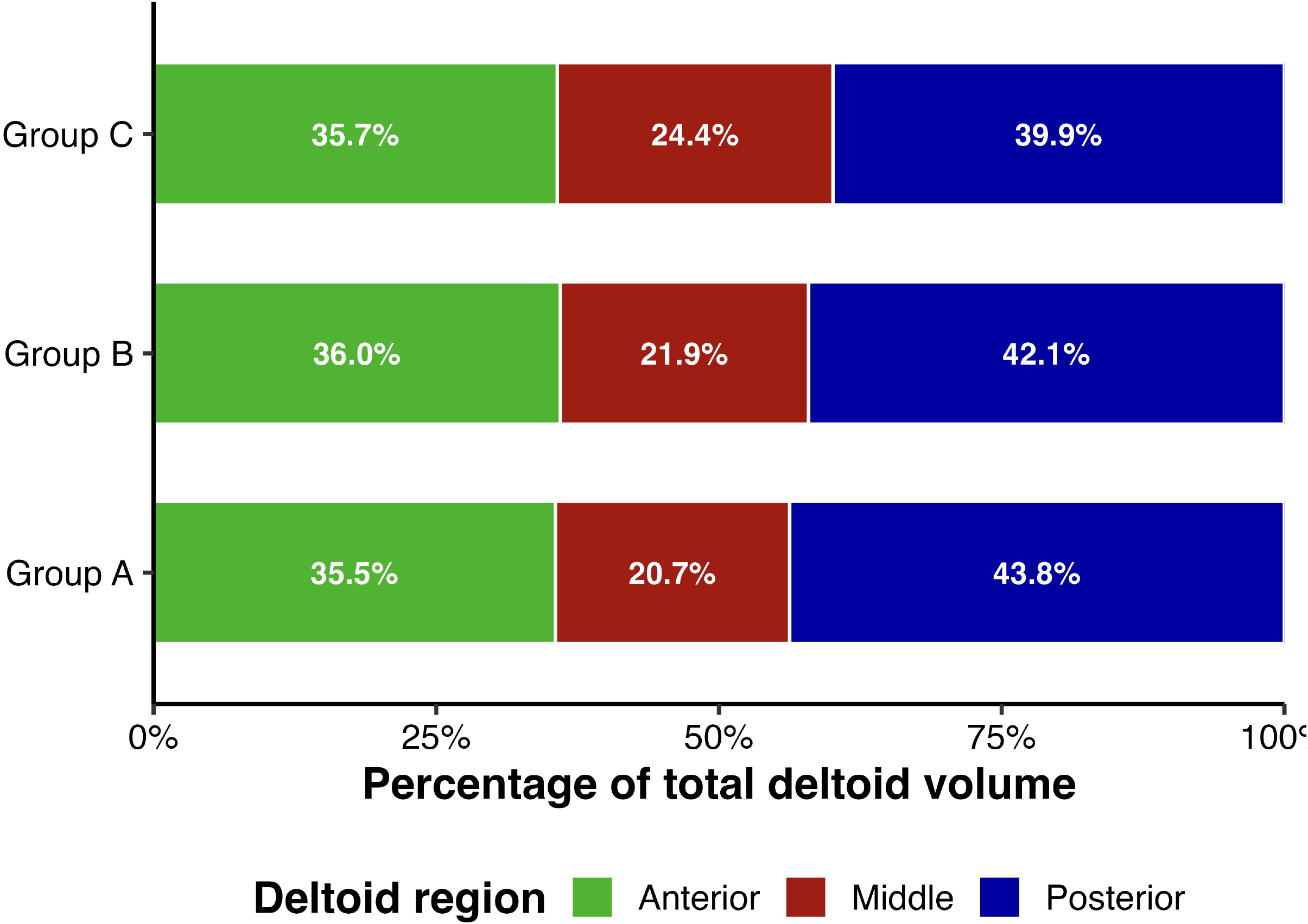
Deltoid muscle via subgroup composition by vaccine-eligibility group (Group A 65+ without risk conditions, Group B 65+ with risk conditions and Group C <65 with risk conditions). Bars show the mean anterior, middle and posterior subgroup volumes as percentages of total bilateral-average deltoid volume. Percentages within each vaccination group sum up to 100%.

The difference was 30.66 ml (29.04 - 32.29) in women and 79.72 ml (78.10 - 81.33) in men. It was 52.02 ml (50.75 - 53.28) on the left and 58.36 ml (57.09 - 59.63) on the right (***Table 4, Figure 3***). The right-left difference was 6.35 ml (5.39 - 7.30, p<0.001), averaged across vaccination-eligibility groups. All pairwise group comparisons remained significant after Tukey adjustment (all p<0.001, ***Table S1***).

### Discussion

With increasing vaccine administration, the incidence of SIRVA has risen [4], with an annual incidence of 1.5% - 2.5% following inactivated influenza vaccination [24]. Accordingly, establishing an evidence-based technique for IM deltoid vaccine administration could help reduce SIRVA. Herein, we quantified deltoid muscle volume in adults eligible for UKNFIP. Using a deep-learning automated segmentation model, we found that posterior deltoid volume was greater than middle deltoid volume on average. The size of this difference varied by vaccination-eligibility group, sex, and side. Greater muscle volume may potentially indicate greater available muscle bulk for intramuscular injection; on this basis, the posterior deltoid may offer the largest volumetric advantage over the middle deltoid in Group A. However, volumetric differences alone are insufficient to establish a preferred vaccination site or recommend a change in current practice.

Dixon MRI enables quantitative evaluation of the shoulder musculature, including deltoid and rotator cuff muscles, providing volumetric and relative fat-fraction measurements [25-28]. MRI has been used in healthy adults by Holzbaur et al. to measure upper limb muscle volumes [29], as well as in patients undergoing arthroscopic rotator cuff repair by Turkmen et al., who measured deltoid muscle volume pre- and post-operatively [30]. In our study, we used Dixon MRI to compare the middle and posterior deltoid volumes in a vaccine-eligible population. Other studies have employed CT-based approaches. Mardourian et al. [12] utilised CT to evaluate the distance from the skin to the deltoid fascia and deltoid muscle width across specific trajectories to determine the ideal IM deltoid injection site. Vervaecke et al. [31] used quantitative three-dimensional CT imaging to quantify sex- and age-based alterations in deltoid in healthy adults. Rajabzadeh-Oghaz et al. [32] developed a transformer-based deep learning model using CT images for automated shoulder muscle segmentation and quantification. In a retrospective review of McClatchy et al. [33] in patients following reverse total shoulder arthroplasty, CT or MRI were utilised to investigate muscle volume. Although less accurate and more operator-dependent than CT [12], earlier studies employed ultrasonography (US) for volumetric purposes [6, 34]. In line with this, Nakajima et al. [6] used US to measure the thickness of subcutaneous tissue and deltoid muscle to establish a safer IM injection site at the intersection points between the anteroposterior axillary line and the perpendicular line from the mid-acromion.

Despite the importance of accurate landmarking in preventing SIRVA, consensus on the safest IM deltoid injection site remains lacking. Previous studies suggested that a safe IM injection site corresponds to a point approximately 5 to 9 centimetres below the acromion process within the middle deltoid part [7, 35]. Nonetheless, Cook et al. [8] reported that these conventional sites have the potential to cause neurovascular complications, including damage to the axillary nerve or its branches, highlighting the importance of identifying alternate injection sites. Recently, Kodaka et al. [36] used US to measure the distances from the conventional injection site to the axillary nerve, radial nerve, and subdeltoid bursa. They proposed an injection site 3.0-3.5 cm below the axillary horizontal line as a safer point for injection. Nonetheless, landmark-based recommendations may not reliably correspond to anatomically safe zones because of substantial inter-individual anatomical variation [36]. Mardourian et al. [12] proposed a point 4 cm distal to and in line with the posterolateral corner of the acromion.

Volumetric examination may represent a more comprehensive descriptor of muscle architecture and morphology compared to single thickness- or part-based measurements [37]. Unlike previous studies which investigated deltoid muscle thickness [6, 8-10] or subcutaneous fat pad [12], our study utilised volumetric analysis of the deltoid muscle. The posterior deltoid had the largest average volume across all groups and in both sexes. The difference between middle and posterior deltoid volumes varied between at-risk vaccination groups, suggesting an age-related correlation between the deltoid muscle parts requiring further investigation outside the scope of this publication. However, this work supports the posterior deltoid as an alternative deltoid IM injection site which may be more relevant in the over-65s where the difference in volume, with the middle deltoid, appears larger in both men and women. A major strength of this study is its large sample size derived from UK Biobank neck-to-knee MRI data, and including participants from the three UK NFIP eligibility groups. We also utilised Dixon MRI for detailed volumetric assessment and a deep learning model to automate muscle segmentation.

This study has several limitations. Participants were selected using the inclusion criteria for the UKNFIP; therefore, younger healthy adults were not included. Individuals with shoulder disorders or anthropometric variations that may influence deltoid anatomy were also not specifically evaluated. A significant majority of participants in our study were of white ethnicity, which was not representative of the UK population. Thirdly, a small group (33 men and 61 women) were excluded due to the limited field of view of their scans. Collectively, these may limit the generalisability of our findings to other ethnic and demographic populations. Finally, needle length and the proximity of proposed injection sites to tendons, nerves, blood vessels, and other non-target structures were not assessed. Future studies should investigate these anatomical relationships, simulate a landmark-based approach and consider other shoulder-affecting pathologies to validate the safety of the proposed injection site.

### Conclusion

Using automated segmentation, we quantified anterior, middle, and posterior deltoid muscle volumes in a pilot UK Biobank cohort. Posterior deltoid volume was greater than middle deltoid volume, on average, and the difference varied by vaccination-eligibility group, sex, and side. The posterior deltoid may therefore be considered for further anatomical assessment as a possible IM injection target.

## Abbreviations

SIRVA: Shoulder injury related to vaccine administration
IM: intramuscular
ML: machine learning
UKNFIP: United Kingdom National Flu Immunisation Programme.

## Declarations

### Data availability

The data that support the findings of this study are available from the UK Biobank (https://www.ukbiobank.ac.uk), but restrictions apply to the availability of these data, which were used under license for the current study, and so are not publicly available. Data are, however, returned by us to the UK Biobank where they will be fully available on request.

### Author contributions

D.A., E.L.T., B.W., and N.B. conceived the study. D.A., J.D.B., N.B., B.W., E.L.T., and M.N. designed the study. N.B. and B.W. implemented the methods. D.A. and M.V. performed the deltoid annotations. B.W. and N.B. developed the deep learning segmentation models. D.A. and A.D. assessed the segmentation quality. N.B. and B.W. performed the image analysis. M.N. performed the data analysis, defined the disease and physiological condition categories, and performed the statistical analysis. F.D., M.N., N.B., B.W., and D.A. drafted the manuscript. All authors read and approved the manuscript.

### Author Approval

All authors have seen and approved the manuscript.

### Ethical Approval

Institutional Review Board approval was obtained (REC reference: 11/NW/0382).

## Acknowledgments

This research has been conducted using the UK Biobank Resource under Application Number 23889.

We thank the participants in the UK Biobank imaging study.

