## Supplementary figures and tables for "Volumetric Analysis of the Deltoid Muscle From Neck-to-Knee Dixon MRI: A pilot population assessment of a posterior intramuscular injection target"

Supplementary material

Figures

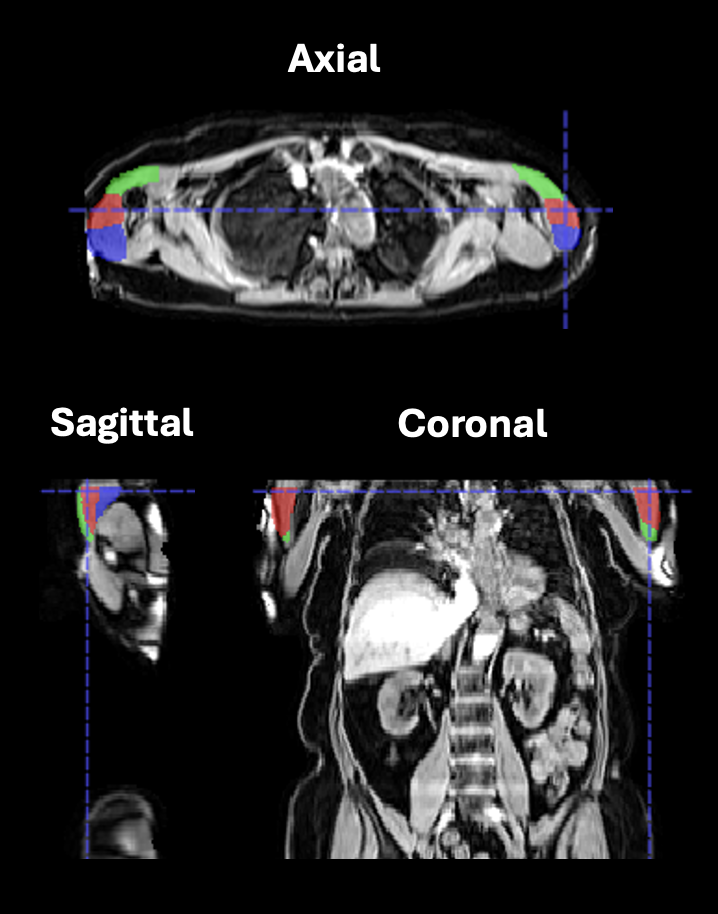

Supplementary Figure 1. Representative dataset of an excluded participant showing field-of-view cut-off using the water-channel image and automated segmentation overlay. (A) Coronal and (B) axial cross-sections with the automated deltoid segmentation masks. Image reproduced by kind permission of UK Biobank ©.

Tables

Supplementary Table 1. List of conditions and the corresponding ICD9 and ICD10 codes used to define vaccine eligibility groups according to the UK influenza programme.

| **Disease** | **Condition** | **ICD9 Codes** | **ICD10 Codes** | **Self-reported** | **OPCs codes** | **Self-reported**  **operation codes** |
| --- | --- | --- | --- | --- | --- | --- |
| Chronic respiratory disease | Chronic bronchitis, emphysema,  COPD,  bronchiectasis,  interstitial lung disease/pulmonary fibrosis,  cystic fibrosis,  pneumoconioses (coal worker's, asbestos, dust-related),  bronchopulmonary dysplasia | 491, 492, 494, 496, 500, 502, 503, 504, 505, 516.3 | E84, J41, J43, J44, J47, J60-J65, J84 | 42017, 131487, 131489, 131491, 131493, 130823, 131501, 131503, 131505, 131507, 131509, 131529 |  |  |
| Cardiovascular disease | Congenital cardiac septal defects,  Tetralogy of Fallot,  coarctation of the aorta,  congenital valve malformations,  great vessel malformations,  heart failure,  atrial fibrillation,  peripheral vascular disease,  pulmonary embolism,  arterial embolism,  venous embolism,  acute myocardial infarction | 410, 411, 415, 416.2, 427, 428, 443, 444, 453, 745, 746,  747.0-747.4 | I21, I22, I23, I26, I48, I50, I73, I74, Q20-Q25 | 131309, 131351, 131355, 131387, 131389, 131401, 132467, 132469, 132471, 132473, 132477 |  |  |
| Chronic or advanced kidney disease | CKD stage 3-5,  end-stage kidney disease,  nephrotic syndrome,  kidney transplantation | 581, 585.3, 585.4, 585.5, 585.6, V42.0 | N04, N18.3, N18.4, N18.5, N18.6, Z94.0 | 132007 | M01, M011, M012, M013, M014, M015, M018, M019 | renal/ kidney transplant_1195 |
| Chronic liver disease | Cirrhosis,  chronic viral hepatitis,  chronic hepatitis,  biliary atresia | 571.2, 571.4, 571.5, 571.6, 751.6 | B18, K73, K74, Q44.2 | 130201, 131665, 131667 |  |  |
| Type 1 diabetes mellitus | Insulin-dependent diabetes mellitus |  | E10 | 130707 |  |  |
| Neurological, neurodevelopmental and  neurodegenerative disorders | Motor neurone disease (ALS),  Parkinson's disease,  multiple sclerosis,  cerebral palsy,  Down syndrome,  learning disability/intellectual disability,  Alzheimer's disease,  vascular dementia,  other dementia | 290, 319, 331.0, 332, 335.2, 340, 758.0 | F01, F02, F03, F81, G12.2, G20, G30, G35, G80, Q90 | 42029, 131023, 131043, 132589, 130837, 130839, 130841, 130843 |  |  |

Supplementary Table 2. Post-hoc comparisons of the posterior-middle deltoid differences. Positive estimates indicate a larger posterior-minus-middle difference in the first-named comparison. Group contrasts are averaged across sides and use Tukey adjustment. The side contrast is averaged across vaccination groups.

| Comparison | Difference (95% CI), ml | Adjusted p |
| --- | --- | --- |
| A − B | 8.58 (5.65 - 11.51) | <0.001 |
| A − C | 17.73 (14.80 - 20.67) | <0.001 |
| B − C | 9.15 (6.21 - 12.09) | <0.001 |
| Men − women | 49.05 (47.05 - 51.06) | <0.001 |
| Right − Left | 6.35 (5.39 - 7.30) | <0.001 |
