## Supplementary material for "Volumetric Analysis of the Deltoid Muscle From Neck-to-Knee Dixon MRI: A pilot population assessment of a posterior intramuscular injection target": Tables

| Characteristic | **Overall** n = 5,239 | **Group A** n = 1,760 | **Group B** n = 1,744 | **Group C** n = 1,735 |
| --- | --- | --- | --- | --- |
| **Sex** |  |  |  |  |
| Women | 2,606 (49.7%) | 880 (50.0%) | 870 (49.9%) | 856 (49.3%) |
| Men | 2,633 (50.3%) | 880 (50.0%) | 874 (50.1%) | 879 (50.7%) |
| **Ethnicity** |  |  |  |  |
| White | 5,112 (97.7%) | 1,729 (98.4%) | 1,716 (98.5%) | 1,667 (96.2%) |
| Asian | 30 (0.6%) | 10 (0.6%) | 5 (0.3%) | 15 (0.9%) |
| Black | 32 (0.6%) | 4 (0.2%) | 6 (0.3%) | 22 (1.3%) |
| Chinese | 16 (0.3%) | 4 (0.2%) | 3 (0.2%) | 9 (0.5%) |
| Other ethnic group | 42 (0.8%) | 11 (0.6%) | 12 (0.7%) | 19 (1.1%) |
| Age (years) | 66.67 ± 7.22 | 70.40 ± 3.78 | 71.35 ± 3.96 | 58.18 ± 4.35 |
| Townsend deprivation index | -1.75 ± 2.81 | -2.02 ± 2.66 | -1.80 ± 2.75 | -1.42 ± 2.96 |
| Height (cm) | 169.08 ± 9.34 | 168.12 ± 9.23 | 168.48 ± 9.22 | 170.66 ± 9.37 |
| Weight (kg) | 78.93 ± 18.15 | 73.90 ± 13.77 | 77.94 ± 16.32 | 85.02 ± 21.71 |
| Body mass index (kg/m²) | 27.54 ± 5.70 | 26.06 ± 3.87 | 27.40 ± 5.13 | 29.19 ± 7.19 |
| Waist circumference (cm) | 91.39 ± 14.64 | 88.18 ± 11.78 | 91.92 ± 13.91 | 94.06 ± 17.11 |
| Hip circumference (cm) | 102.74 ± 10.94 | 99.99 ± 7.92 | 102.68 ± 10.07 | 105.53 ± 13.38 |
| Waist-to-hip ratio | 0.89 ± 0.09 | 0.88 ± 0.09 | 0.89 ± 0.09 | 0.89 ± 0.10 |
| Dominant-hand grip strength (kg) | 30.35 ± 10.43 | 29.45 ± 9.79 | 28.84 ± 9.94 | 32.75 ± 11.09 |
| Moderate-to-vigorous activity (MET-hours/week) | 18.74 ± 13.70 | 20.45 ± 13.42 | 18.74 ± 13.46 | 17.08 ± 13.99 |
| Total muscle volume (L) | 17.86 ± 4.63 | 17.18 ± 4.32 | 17.28 ± 4.27 | 19.14 ± 4.99 |
| Iliopsoas muscle volume (ml) | 636.79 ± 169.21 | 615.28 ± 156.43 | 614.99 ± 157.53 | 680.51 ± 184.00 |
| Values are n (%) or mean ± SD. | | | | |

Table 1.

Table 2.

| Compartment | Likert score, mean ± SD | Weighted kappa (95% CI) | Exact agreement |
| --- | --- | --- | --- |
| Overall | 4.27 ± 0.73 | 0.583 (0.452 - 0.713) | 62.5% |
| Left Anterior | 4.40 ± 0.67 | 0.676 (0.397 - 0.954) | 70.0% |
| Left Middle | 4.28 ± 0.82 | 0.580 (0.256 - 0.904) | 60.0% |
| Left Posterior | 4.10 ± 0.74 | 0.630 (0.387 - 0.873) | 60.0% |
| Right Anterior | 4.65 ± 0.48 | 0.158 (-0.241 - 0.557) | 60.0% |
| Right Middle | 4.30 ± 0.72 | 0.519 (0.175 - 0.864) | 65.0% |
| Right Posterior | 3.88 ± 0.72 | 0.461 (0.054 - 0.867) | 60.0% |

Table 3.

|  |  | Sex | | Vaccination-eligibility group | | | Side | |
| --- | --- | --- | --- | --- | --- | --- | --- | --- |
| Deltoid parts | **Overall**  n = 5,239 | **Women**  n = 2,606 | **Men**  n = 2,633 | **Group A**  n = 1,760 | **Group B**  n = 1,744 | **Group C**  n = 1,735 | **Left**  n = 5,239 | **Right**  n = 5,239 |
| Total deltoid | 260.99 ± 59.76 | 214.35 ± 33.88 | 307.16 ± 41.04 | 263.36 ± 58.36 | 255.37 ± 56.59 | 264.25 ± 63.76 | 246.14 ± 68.87 | 275.85 ± 72.30 |
| Anterior | 92.59 ± 19.78 | 77.89 ± 12.58 | 107.14 ± 14.00 | 92.86 ± 18.66 | 91.20 ± 18.78 | 93.71 ± 21.70 | 89.62 ± 20.86 | 95.56 ± 21.84 |
| Middle | 56.52 ± 17.69 | 52.86 ± 15.71 | 60.15 ± 18.76 | 53.27 ± 14.06 | 54.37 ± 15.54 | 61.99 ± 21.37 | 52.17 ± 19.13 | 60.88 ± 25.37 |
| Posterior | 111.88 ± 39.83 | 83.60 ± 24.04 | 139.87 ± 31.78 | 117.23 ± 37.42 | 109.80 ± 37.92 | 108.55 ± 43.39 | 104.35 ± 42.53 | 119.41 ± 47.35 |
| Values are mean ± SD in ml. | | | | | | | | |

| Comparison | Adjusted posterior − middle difference (ml)  mean (95% CI) | p-value |
| --- | --- | --- |
| Vaccination-eligibility group | | |
| Overall test |  | <0.001 |
| Group A | 63.96 (61.85 - 66.07) |  |
| Group B | 55.38 (53.26 - 57.50) |  |
| Group C | 46.23 (44.10 - 48.35) |  |
| Sex | | |
| Overall test |  | <0.001 |
| Women | 30.66 (29.04 - 32.29) |  |
| Men | 79.72 (78.10 - 81.33) |  |
| Side | | |
| Overall test |  | <0.001 |
| Left | 52.02 (50.75 - 53.28) |  |
| Right | 58.36 (57.09 - 59.63) |  |
| All estimates are adjusted for other model terms. | | |

Table 4.
